# Antibody and memory B cell responses to the dengue virus NS1 antigen in individuals with varying severity of past infection

**DOI:** 10.1101/2022.08.30.22279380

**Authors:** Shyrar Tanussiya Ramu, Madushika Dissanayake, Chandima Jeewandara, Farha Bary, Michael Harvie, Laksiri Gomes, Ayesha Wijesinghe, Graham S. Ogg, Gathsaurie Neelika Malavige

**Affiliations:** Allergy Immunology Cell Biology Unit, Department of Immunology and Molecular Medicine, University of Sri Jayewardenepura, Sri Lanka; MRC Human Immunology Unit, MRC Weatherall Institute of Molecular Medicine, University of Oxford, Oxford, United Kingdom

**Author notes:** **Correspondence should be addressed to:** Prof. Neelika Malavige DPhil (Oxon), FRCP (Lond), FRCPath (UK), AICBU, Department of Immunology and Molecular Medicine, Faculty of Medical Sciences, University of Sri Jayewardenepura, Sri Lanka.

## Abstract

**Background:** To further understand the role of NS1 specific antibodies (Abs) in disease pathogenesis, we compared neutralizing antibody levels (Nabs), NS1-Ab levels, IgG antibody sub-class profiles and NS1 specific memory B cell responses (Bmems) in individuals, with varying severity of past dengue.

**Methods:** Nabs (Neut50 titres) were assessed using Foci Reduction Neutralization Test (FRNT) and in-house ELISAs were used to assess NS1-Abs and NS1-Ab subclasses for all four DENV serotypes in individuals with past DF (n=22), those with past DHF (n=14) and seronegative (SN) individuals (n=7). B cell ELISpot assays were used to assess NS1-specific Bmem responses.

**Results:** 15/22 (68.18%) individuals with past DF and 9/14 (64.29%) individuals with past DHF had heterotypic infections. Neut50 titres were found to be significantly higher for DENV1 than DENV2 (p=0.0006) and DENV4 (p= 0.0127), in those with past DHF, whereas there was no significant difference in titres was seen for different DENV serotypes in those with past DF. Overall NS1-Ab to all serotypes and NS1-specific IgG1 responses for DENV1, 2 and 4 serotypes were significantly higher in those with past DHF than individuals with past DF. Those with past DHF also had higher IgG1 than IgG3 for DENV1 and DENV3, whereas no differences were seen in those with past DF. Over 50% of those with past DF or DHF had NS1-specific Bmem responses to >2 DENV serotypes. There was no difference in the frequency of Bmem responses to any of the DENV serotypes between individuals with past DF and DHF. Although the frequency of Bmem responses to DENV1 corelated with DENV1 specific NS1-Abs levels (Spearman r=0.35, p=0.02), there was no correlation with other DENV serotypes.

**Conclusions:** We found that those with past DF had broadly cross reactive Nabs, while those with past DHF had higher NS1-Ab responses possibly with a different functionality profile than those with past DF. Therefore, it would be important to further evaluate the functionality of NS1-specific antibody and Bmem responses to find out the type of antibody repertoire that is associated with protection against severe disease.

## Introduction

Infections due to the dengue virus (DENV) is the leading mosquito borne viral infection globally, and due to the global burden of infection, the World Health Organization named it as one of the top ten threats to global health in year 2019 [1]. The age-standardized incidence rates, mortality rates and disability adjusted life years (DALYs) have risen markedly from year 1990 to year 2017 [2]. For instance, there has been 107.6% global increase in DALYs from year 1990 to year 2017 [2]. As there is no specific treatment for dengue, currently all patients who present with dengue fever (DF) are carefully monitored for early detection of complications for timely interventions.

Although the majority of individuals infected with the DENV develop asymptomatic or mild illness, a proportion of individuals develop dengue haemorrhagic fever (DHF), which is characterized by plasma leakage [3]. Endothelial dysfunction, which leads to plasma leakage can be due to many factors such as DENV NS1 antigen, an impaired and dysfunctional antiviral response, antibodies specific to the previous DENV serotype enhancing infection, mast cell degranulation and an aberrant response by monocytes [4; 5; 6]. Of these many factors that cause endothelial dysfunction, the NS1 is emerging as one of the most important factors that lead to endothelial dysfunction. NS1 is secreted from infected cells and circulates as a barrel shaped hexamer, with a lipid core in the middle [7]. NS1 has shown to directly disrupt the endothelial glycocalyx layer, cause disassembly of the intercellular gap junctions, activate immune cells to produce proinflammatory cytokines, activate complement, activate monocytes to produce prostaglandins and inflammatory phospholipase A2 enzymes and also induce production of immunosuppressive cytokines [4; 8; 9; 10; 11]. It has been shown that high levels of NS1 during early illness is associated with progression to severe disease in some studies, although the NS1 levels and positivity rates during early illness have shown to change with the infecting DENV serotype [12; 13; 14].

While most individuals infected with the DENV do not develop severe dengue, secondary dengue infections, where an individual is infected with a subsequent different serotype than the initial infection, has shown to be an important risk factor for severe illness [15; 16]. Severe disease during secondary dengue is thought in many cases to be associated with antibody dependent enhancement (ADE) where poorly neutralizing antibodies specific to the previous DENV serotype, facilitating infection into FcγR bearing immune cells, and thereby facilitate infection [17; 18]. Although NS1 is known to contribute to severe disease by many mechanisms, the NS1 antigen levels are lower in patients with secondary dengue, than in those with primary dengue [19; 20], possibly due to the lower viral loads [21]. Therefore, in addition to the direct pathogenic effects of NS1 antigen, NS1 antigen-antibody complexes may also play a role in disease pathogenesis. NS1 specific antibodies have shown to cause disease pathogenesis by cross reacting with many different host-proteins such as endothelial cells, fibrinogen and platelets [22]. NS1-Abs have shown to bind to the endothelium resulting in expression of ICAM-1 and release of many proinflammatory cytokines and have shown to contribute to liver damage in murine models [22]. NS1-Abs specific for the amino acid residues 311–330 of NS1 antigen, have shown to bind to protein disulfide isomerase on platelets, thereby contributing to thrombocytopenia [23]. In contrast, NS1-Ab have also shown to be protective in murine models and it has shown that that functionality of the NS-Abs are important in protection [24].

We previously showed that the NS1 specific antibody levels significantly increased in those who progressed to develop DHF compared to those with DF in secondary dengue infections [25]. We also showed that those with acute DF had a higher frequency of antibody responses to different regions of NS1 than in those with DHF [25]. It was recently shown that individuals who subsequently developed inapparent dengue due to DENV3 had higher baseline levels of antibodies to NS1 and envelope protein with more functional antibody responses [24]. Therefore, the antibody levels to NS1, the levels of neutralizing antibodies, cross reactivity of antibodies and memory B cell responses, all could contribute to disease severity or protection. In this study, we compared to neutralizing antibody levels to different DENV serotypes, NS1-Ab levels, the IgG antibody sub-class profiles and NS1 specific memory B cell responses in healthy individuals, with varying severity of past dengue, to further understand the factors that lead to severe disease in dengue.

## Materials and methods

### Study participant recruitment and sample collection

Healthy adult individuals (n=43) were recruited for the study following informed consent. Individuals who had been previously hospitalized and had been diagnosed as having DHF due to the presence of pleural effusions or ascites with platelet counts <100,000/cells^3^ based on WHO 2011 guidelines [26], were considered to have past DHF (n=14). Those who were hospitalized but did not have DHF, were considered to have DF (n=11) and those who were seropositive for the DENV but never hospitalized for a febrile illness were considered to have inapparent dengue/undifferentiated DF (n=11). Individuals who had never been hospitalized for a febrile illness and were seronegative for the DENV by ELISA, were considered as DENV naïve individuals (n=7).

Ethical approval was obtained from the Ethics Review Committee of the Faculty of Medical Sciences, University of Sri Jayewardenepura, Sri Lanka.

#### ELISA to determine serostatus to the DENV

Panbio Dengue Indirect IgG ELISA (Panbio, Brisbane, Australia,) was carried out to identify the seropositivity to the DENV. The assay was carried out according to the manufacturer’s instructions and accordingly, those who had PanBio units (indirect measure of DENV IgG) of >11 were considered positive, 2 to 11 was considered equivocal and <2 was considered negative as previously proposed in a research carried out by Lopez et al. [27], in order to eliminate false-negatives. Based on these interpretations, 11/18 were classified as seropositive and the remaining 7 were considered seronegative individuals.

### Foci Reduction Neutralization Test (FRNT) to determine neutralizing antibody titres

Neutralizing antibody titres to DENV1 WestPac74, DENV2 S16803, DENV3 CH53489 and DENV4 TVP-376 (donated by Prof. Aravinda de Silva) were assessed using FRNTs as previously described [28]. The viruses were propagated in C6/36 cell lines, after which the foci forming assays (FFAs) were carried out on Vero-81 cells to determine the virus concentration in FFU/ml (foci forming units/ml). These viruses were used to assess the neutralizing antibodies using FRNTs. Vero-81 cells were seeded in 96-well tissue culture treated plates and incubated overnight. The patient sera were diluted at a four-point dilution series of 1:10, 1:40, 1:160 and 1:640 in DMEM (Gibco, Life Technologies, USA) supplemented with 2% FBS and was incubated for an hour with the calculated amount of each DENV separately. The diluted serum and virus mix was then added to the Vero-81 cell monolayer after which the plates were incubated for an hour at 37°C with 5% CO_2_. Then overlay media (Carboxymethyl cellulose) was added to the monolayer and the plates incubated at 37°C with 5% CO_2_ for 2 to 3 days. Wells were then fixed with 4% paraformaldehyde (Alfa Aesar, UK) and blocked with blocking buffer (1X perm buffer, Biolegend, USA) with 3% normal goat werum (Sigma, USA). Staining was carried out using a mixture of 4G2 and 2H2 biotinylated antibodies (donated by Prof. Aravinda de Silva). HRP conjugated goat anti-mouse IgG (KPL, SeraCare Life Sciences, USA) was used as the secondary antibody, followed by TrueBlue Peroxidase substrate (KPL, SeraCare Life Sciences, USA). The assays were performed in duplicates.

The RStudio software was used to count the spots and GraphPad Prism version 9 was used to analyse data and to calculate Neut_50_ by plotting neutralization percentages against the log (1/dilution) values. Those with Neut_50_ titres <10 for all 4 serotypes was classified as naïve individuals, those who had Neut_50_ titres ≥10 for only one serotype, or ≥10 for different serotypes but 5 times higher for one serotype only were classified as being monotypic (infected with only one DENV serotype), and those with Neut_50_ titres ≥10 for different serotypes (without higher titres for a single serotype) were considered as having had multitypic infection.

### Determining levels of NS1 specific antibody levels for different DENV serotypes

In order to determine the NS1 antibody levels specific to each serotype of DENV in the sera of these individuals, an in-house indirect ELISA was performed as described previously [25]. Briefly, 96-well plates were coated with DENV1, DENV2, DENV3 and DENV4 NS1 recombinant proteins expressed in mammalian HEK293 cells (Native Antigen, USA), separately. The plates were blocked with blocking buffer buffer (PBS containing 0.05% Tween 20 and 1% Bovine serum albumin (BSA)). The serum samples that were diluted at 1:5000 were added to the plates and the ELISA was developed using goat anti-human IgG biotinylated antibody (Mabtech, Sweden), and Streptavidin Alkaline Phosphatase (Abcam,UK) as the enzyme and Para-nitro-phenyl-phosphatase (PNPP) (Thermofisher Scientific, USA) as the substrate. Absorbance was measured at 405nm using the MPSCREEN MR-96A ELISA reader. The positive cut off threshold was set at mean± SD of the optic density (OD) value of seronegative individuals.

### Detection of NS1 specific IgG1 and IgG3 antibody levels in sera

Levels of NS1 specific IgG1 and IgG3 subclass to NS1 of all 4 DENV serotypes was assessed individually in serum samples using an in-house ELISA. The 96-well ELISA plates was coated at 5µg/ml with DENV1, DENV2, DENV3 and DENV4 NS1 proteins (Native Antigen, USA), separately. The wells were then blocked and serum diluted at 1:250, which was added to the plate. IgG1 and IgG3 levels were detected using biotinylated goat anti-human IgG1 (Mabtech, Sweden, Cat: 3851-6-250) and biotinylated goat anti-human IgG3 (Mabtech, Sweden, Cat: 3853-6-250), respectively. Streptavidin-horse radish peroxidase (Mabtech, Sweden, Cat: 3310-9) and TMB (Mabtech, Sweden, Cat: 3652-F10) substrate was used to develop the reaction. The absorbance values were obtained at 450nm using the MPSCREEN MR-96A ELISA reader.

### Memory B cell ELISpot to determine Bmem responses to DENV NS1 protein

Memory B cell Responses (Bmem) specific for DENV1, DENV2, DENV3 and DENV4 NS1protein was assessed as previously described by us for assessing Bmem responses to different COVID-19 vaccines [29; 30]. Briefly, freshly isolated PBMCs were stimulated in a 24 well plate using IL-2 and R848 (a TLR 7/8 agonist) in RPMI media (Gibco, Life Technologies, USA) supplemented with 10% Fetal Bovine Serum, 1% Penicillin Streptomycin and 1% Glutamine (Gibco, Life Technologies, USA) (R10 media) and incubated at 37 °C with 5% CO_2_ for 3 days. Unstimulated PBMCs were also incubated in R10 media, as a negative control. After an incubation period of 3 days, the cells were rested overnight. The assay was optimized by adding 100,000 cells/well and the optimal coating concentration was determined to be 1µg/ml. 50,000 cells/well were added to the positive control wells. A Human IgG ELISpot BASIC kit (Mabtech 3850-2A) was used according to the manufacturer’s guidelines to quantify IgG-secreting cells specific to DENV NS1 full length recombinant proteins (Native Antigen, UK) for all four serotypes, which was used to coat the wells at 1µg/ml in PBS. Anti-human IgG monoclonal capture antibodies were used to coat the positive control wells and culture media was used for the negative control. The experiments were carried out in duplicates and the spots were counted using the automated AID iSpot Reader System (GmbH Germany). Mean□±□2 SD of the background responses was defined as a positive response.

### Statistical Analysis

Statistical Analysis was carried out using GraphPad Prism version 9. As the data were not normally distributed, non-parametric tests were carried out. All tests were two-tailed. The Mann-Whitney U test was performed to compare differences in antibody levels and the frequency of Bmem between those with past DHF, DF and DENV seronegative individuals. Differences in FRNT responses, NS1-antibody levels and Bmem responses between serotypes were compared using the Friedman test along with a Dunns multiple comparison t test to compare between serotypes. The Wilcoxon’s matched pairs signed rank test was used to compare paired data when responses to IgG1 and IgG3 were compared between the same individuals. Spearman’s rank two-tailed correlation coefficient was used to evaluate the correlation between variables including Bmem responses, antibody levels and Nabs levels (Neut_50_ titres).

## Results

### Neutralizing antibody (Nab) levels in individuals with varying severity of past dengue infections

Out of the 43 healthy individuals that were enrolled in the study, 28/43 (65.1%) were females and 15/43 (34.88%) were males. Their mean age was 34.56 years (±10.24). The Nab levels were assessed in healthy individuals with past DHF (n=14), those with past DF (n=11), those with inapparent dengue/undifferentiated DF (n=11) and in DENV seronegative individuals (n=7). In those with past DHF, the time since illness was a median of 4.5 years (IQR 2.375 to 11.75), in those with DF the time since illness was a median of 6 years (IQR 2 to 9). The time since infection in those with undifferentiated DF was not known, as they did not know when they were infected. Those who were hospitalized due to DF and had undifferentiated DF, were classified together as having past DF (n=22) during the analysis. The Nab levels were expressed as Neut50 titres, indicating the Nabs levels at which 50% neutralization of the virus was achieved.

Of those with past DF, 15/22 (68.18%) had Nabs to several DENV indicating heterotypic infections, 5/22 (22.72%) only had responses to one DENV serotype indicating monotypic infections and 2/22 (9.09%) did not have detectable Nabs despite being seropositive by the ELISA. In those with past DHF, heterotypic infections were seen in 9/14 (64.29%) individuals and monotypic infections in 5/14 (35.71%). In individuals with past DF, 8/22 (36.36%) had Nabs against 4 serotypes (Figure 1A), with 15/22 (68.18%) having Nabs against DENV1, 16/22 (72.73%) against DENV2, 15/22 (68.18%) against DENV3 and 12/22 (54.55%) against DENV4. In those who had past DHF infections, Nabs against all 4 serotypes were present in 7/14 (50%) individuals, with all 14/14 (100%) having Nabs against DENV1, 7/14 (50%) having Nabs against DENV2, and 9/14 (64.29%) against DENV3 and DENV4 (Figure 1A). The 7 individuals, who were determined as seronegative for DENV using the IgG Indirect ELISA, were also naïve for neutralizing antibodies against all 4 DENV serotypes.

**Figure 1:**
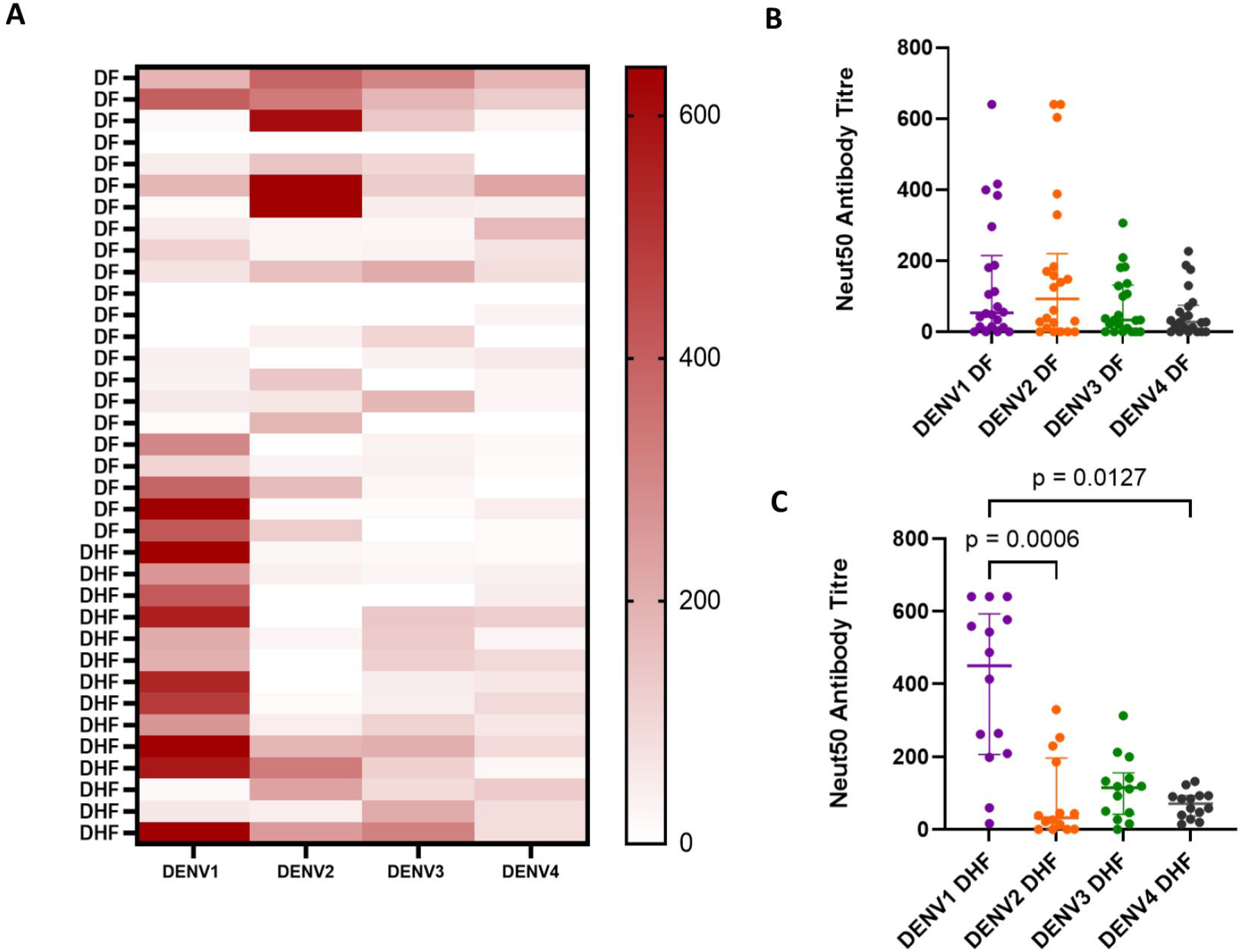
Neutralising antibody levels (Neut50 titres) to DENV1, DENV2, DENV3 and DENV4 in those with varying severity of past dengue infections. Neutralizing antibody (Nab) levels were measured by FRNTs in those who had past DF (n=22) and past DHF (n=11), and the Nab levels for each individual for each DENV serotype is shown in the heat map (A). The differences in the Nab (Neut50 titres) between different DENV serotypes were analysed in those with past DF (B) and past DHF (C) using the Friedman test. All tests were two sided. The lines represent the median and the interquartile range.

The Nabs levels (Neut50 titres) were found to be significantly higher for DENV1 than DENV2 (p=0.0006) and DENV4 (p= 0.0127), in those with past DHF, whereas there was no significant difference between the Neut50 titres for different DENV serotypes in those with past DF (Figure 1B and C). The Nabs for DENV1 (p=0.0006) and DENV4 (p= 0.0436) were significantly higher in those with past DHF compared to those with past DF (Figure 2). Although not significant (p=0.4241), the Neut50 titres were higher for DENV2 (median 93.1, IQR 8.07 to 220.1) in those with past DF compared to past DHF (median 33.42, IQR 0.94 to 196.5).

**Figure 2:**
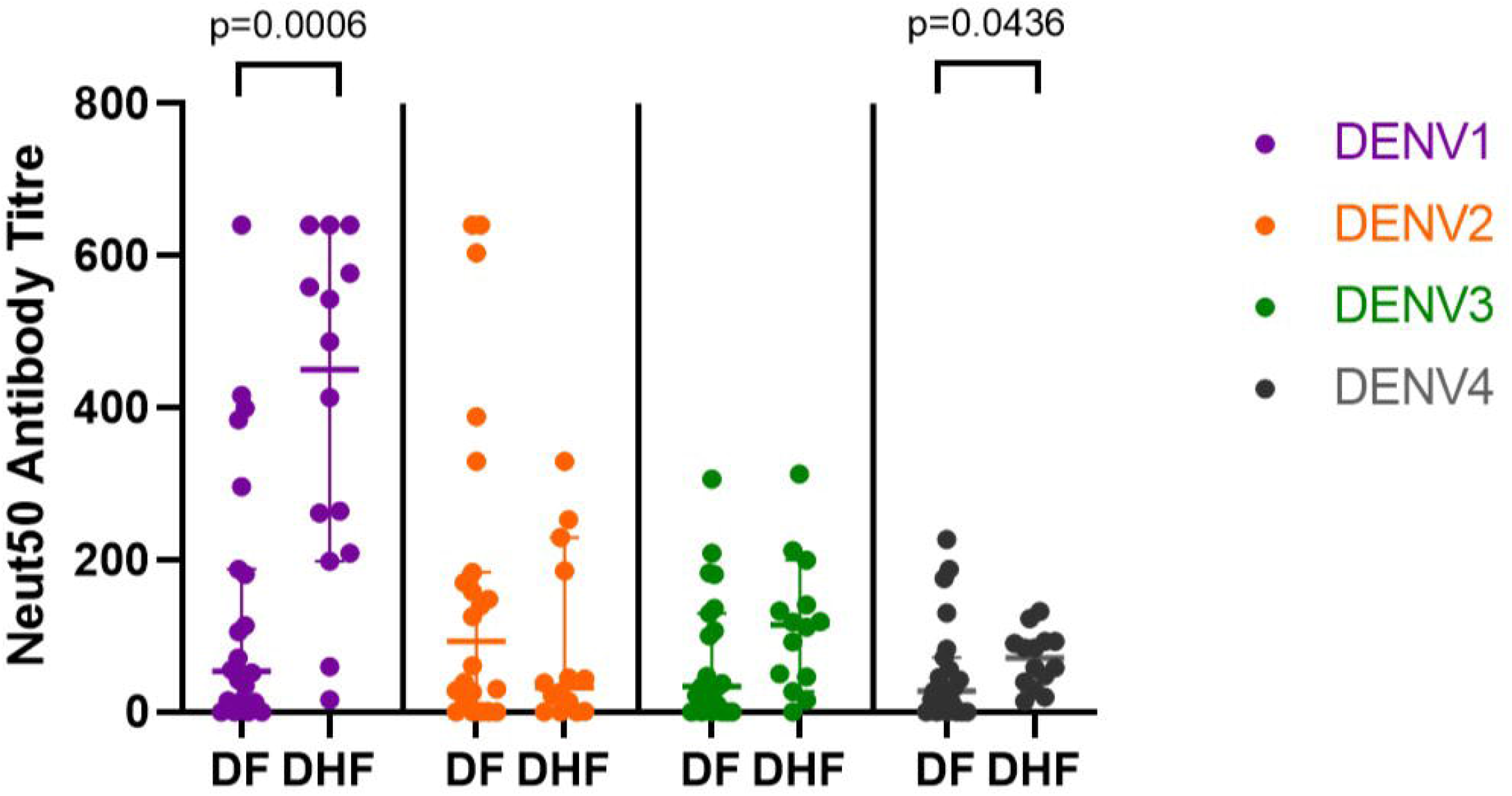
Comparison between neutralising antibody levels (Neut50 titres) for different DENV serotypes of those with varying severity of past DENV in each serotype. The differences in the Nab levels (Neut50 titres) between those with past DF (n=22) and past DHF (n=14) were analysed using the Mann-Whitney U test for the different DENV serotypes. All tests were two-tailed. All tests were two sided. The lines represent the median and the and the interquartile range.

### Comparison of serotype specific NS1 antibody responses in those with varying severity of past dengue

We previously showed that NS1-Ab responses were significantly higher in those with past severe dengue, but we had only investigated NS1-Ab responses to one DENV serotype [25]. Therefore, to determine the relationship between NS1-Ab responses to different serotypes and the extent of cross reactivity, we assessed the NS1-Ab levels to all four serotypes in individuals with past DHF or DF and in seronegative individuals. Both individuals with past DF and past DHF had the highest antibody levels for NS1 of DENV2. In those with past DF, Ab levels against DENV2 NS1 were significantly higher than DENV1 (p=0.0097), DENV3 (p<0.0001) and DENV4 (p<0.0001) (Figure 3A). A similar pattern was seen in those with past DHF, where NS1-Ab levels against DENV2 were significantly higher than DENV3 (p<0.0001) and DENV4 (p<0.0001), and NS1-Ab levels against DENV1 was significantly higher than DENV4 (p=0.0127) (Figure 3B).

**Figure 3:**
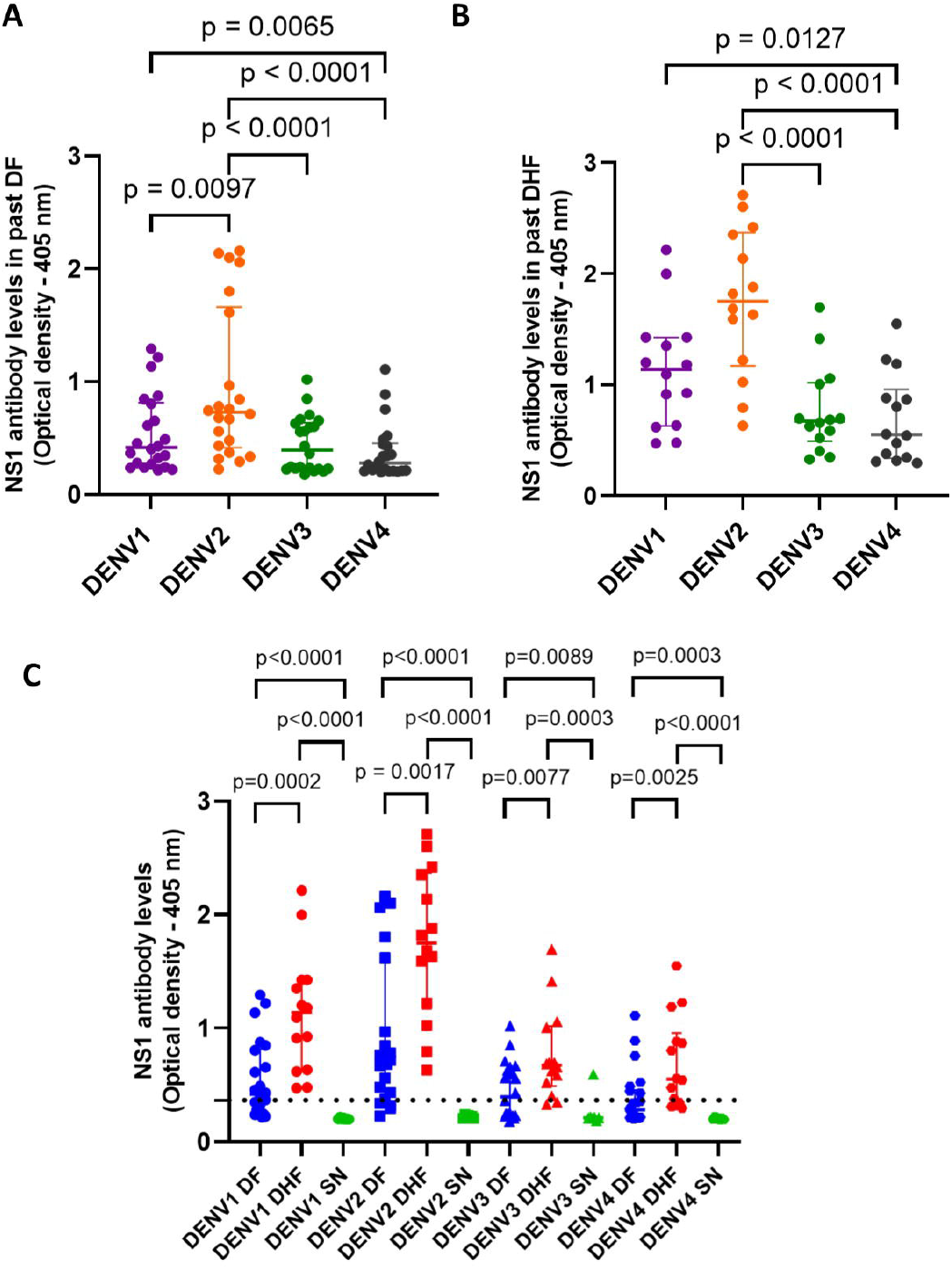
NS1-specific antibody responses to the four DENV serotypes in those with varying severity of past dengue infections. NS1-specific antibodies to the DENV serotypes were assessed by an in-house ELISA in those with past DF (n=22), those with past DHF (n=14) and seronegative individuals (n=7). Comparison of NS1-Ab responses between the serotypes in those with past DF (A) and past DHF (B) were analysed using the Friedman test. The differences between NS1-specific antibody responses in individuals with past DF, past DHF and seronegative (SN) individuals were analysed using the Mann Whitney U test (C). All tests were two tailed. The horizontal dotted line represents the positive cut-off (mean□±□2 SD of the background responses). The lines represent the median and the interquartile range.

NS-1Ab responses in those with past DHF were significantly higher than individuals with past DF for all four DENV serotypes (DENV1, p=0.0002; DENV2, p=0.0017; DENV3, p=0.008 and DENV4, p=0.003) (Figure 3C). DENV3 specific NS1-Ab responses were seen in one seronegative individual (Figure 3C), although this individual did not have any Nabs to any of the DENV serotypes. The NS1-Ab responses positively and significantly correlated with the neutralizing antibody levels (Neut50 titres in all individuals (n=43) to DENV1 (Spearman r=0.69, p<0.0001), DENV2 (Spearman r=0.53, p=0.0003), DENV3 (Spearman r=0.62, p<0.0001) and DENV4 (Spearman r=0.68, p<0.0001) (Supplementary figure 1).

### Comparison of NS1 IgG1 and IgG3 subclass specific antibodies in those with varying severity of past dengue

We previously reported that NS1-Abs of patients with acute DF predominantly recognised different regions of the protein than those with acute DHF [25]. However, apart from the specificity of NS1-Abs and quantitative differences of NS1-Abs in individuals with varying severity of illness, the functionality of NS1-Abs could differ, thereby contributing to disease pathogenesis. As the Fc portion of an antibody is shown to be critical in immune complex formation, inducing antibody dependent cell mediated cytotoxicity (ADCC) by NK cells, ability to activate phagocytes and complement [31; 32], we assessed the levels of NS1-specific IgG1 and IgG3 responses for all four DENV serotype in our cohort.

Those with past DHF had significantly higher NS1-specific IgG1 responses to DENV1 (p=0.0008), DENV2 (p=0.0013) and DENV4 (p=0.0328) than in those with past DF (Figure 4A). Those with past DHF also had higher NS1-IgG3 antibody responses to DENV1 (p=0.0011) and DENV4 (p=0.0165) compared to those with past DF and seronegative individuals (Figure 4B). Individuals with past DHF had significantly higher NS1-IgG1 Ab responses than NS1 IgG3 for DENV1 and DENV3 although this difference was only significant for DENV3 (p=0.038) but not for any other DENV serotypes. (Figure 4C-F). There was no difference between the NS1-IgG3 Ab responses and NS1-IgG1 Ab responses in those with past DF.

**Figure 4:**
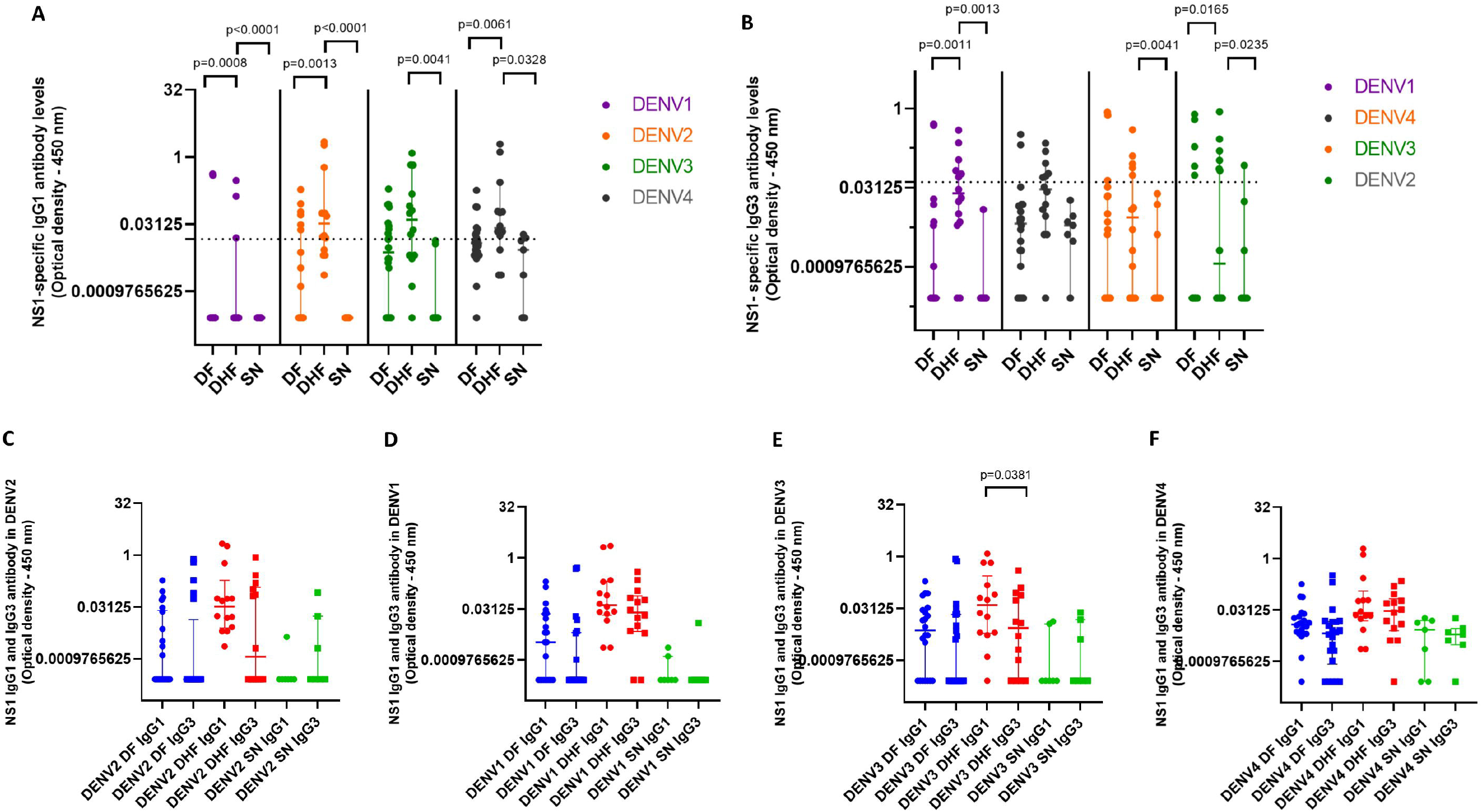
NS1-specific IgG1 and IgG3 responses to the four DENV serotypes in those with varying severity of past dengue infections. NS1-specific IgG1 and IgG3 antibody levels were measured for the four DENV serotypes in individuals with past DF (n=22), those with past DHF (n=14) and seronegative (SN) individuals (n=7) using an in-house ELISA. Comparison in IgG1 responses (A) and IgG3 responses (B) between those with past DF, DHF and seronegative individuals were analysed using the Mann Whitney U test. The horizontal dotted line represents the positive cut-off (mean□±□2 SD of the background responses). Comparison between IgG1 and IgG3 responses in those with past DF, past DHF and seronegative individuals to DENV1 (C), DENV2 (D), DENV3 (E) and DENV4 (F) were analysed using the Wilcoxon Signed-rank test. All tests were two tailed. The lines indicate median and the interquartile range.

### Memory B cell (Bmem) responses to NS1 in those with varying severity of past dengue

As Bmem to DENV NS1 has not been assessed before, we sought to investigate the frequency of Bmems in our cohort for different DENV serotypes and the association with NS1-Ab levels and Nabs. Of the 43 individuals, who were assessed for DENV NS1-specific Bmem responses, responses for ≥3 serotypes were seen in 9/22 (40.91%) individuals with past DF and 7/14 (50%) individuals with past DHF (Figure 5A). Of the DENV seronegative individuals, 2/7 (28.57%) had Bmem responses to 2 serotypes (one for DENV1 and DENV4 and the other person to DENV1 and DENV2) while the remaining 5/7 (71.43%) did not response to any of the 4 serotypes. These seronegative individuals did not have any responses to the DENV by the FRNT assay and did not give a positive response for the presence of NS1-Abs to any of the DENV serotypes.

**Figure 5:**
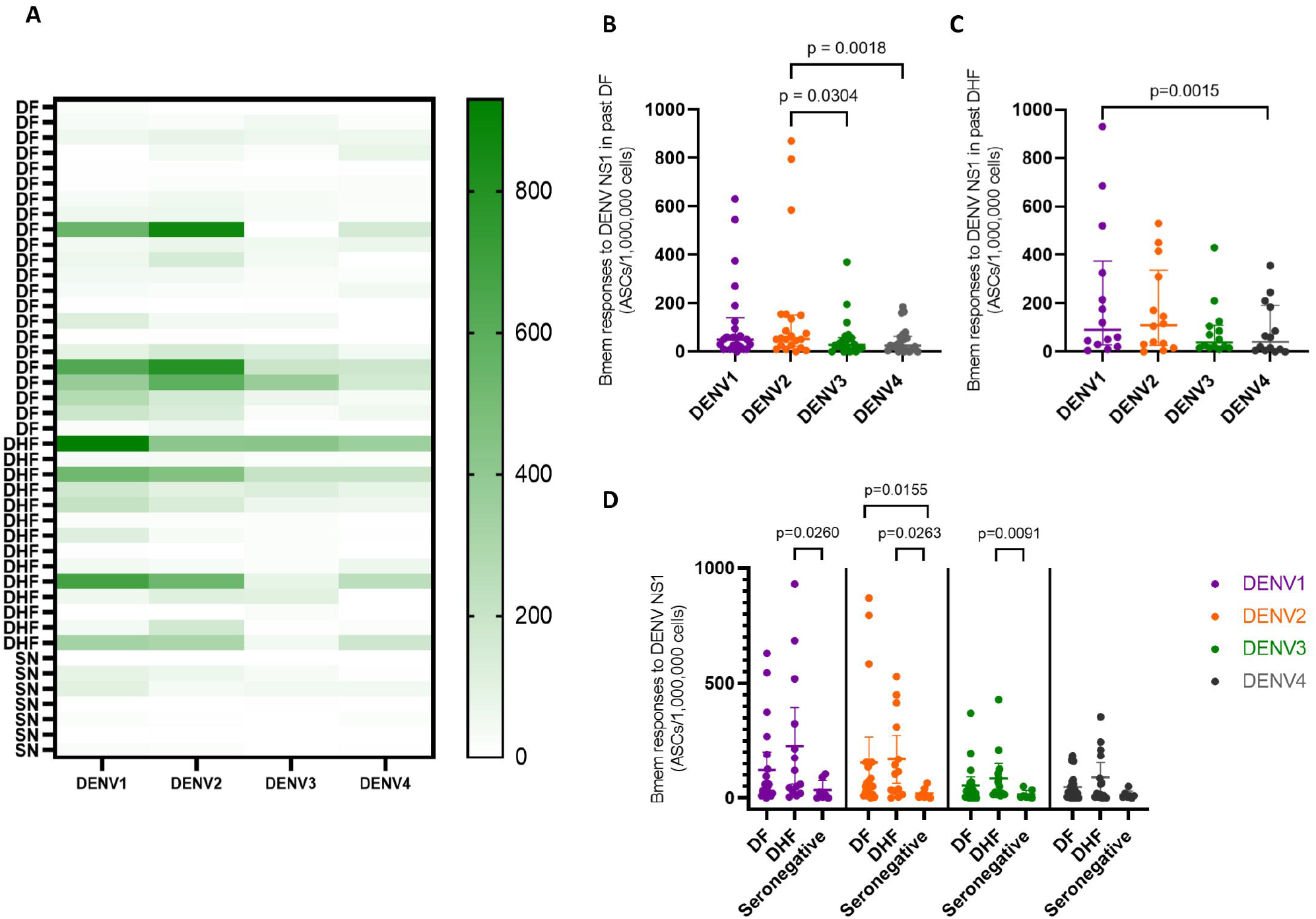
The frequency of NS1-specific memory B cell (Bmem) responses to the four DENV serotypes in individuals with varying severity of past dengue infections. The Bmem responses were measured by B-cell ELISpot assays in those who had past DF (n=22), past DHF (n=11) and seronegative individuals for the four DENV serotypes (SN) (n=7) and are represented in a heat map (A). The differences in the frequency of Bmem responses for four DENV serotypes were analysed in those with past DF (B) and past DHF (C) using the Friedman test. Comparisons in NS1-specific antibody responses between individuals with past DF, past DHF and seronegative (SN) individuals were analysed using the Mann Whitney U test (D). All tests were two tailed. The lines indicate median and the interquartile range.

In those with past DF, the frequency of Bmem responses for DENV2 were significantly higher than DENV3 (p=0.0304) and DENV4 (p=0.018) (Figure 5B), whereas in those with past DHF, Bmem responses were significantly higher against DENV1 than DENV4 (p=0.0015) (Figure 5C). There was no difference in the frequency of Bmem responses to any of the DENV serotypes between individuals with past DF and DHF (Figure 5D).

Although the frequency of Bmem responses to DENV1 showed a positive and significant correlation with DENV1 specific NS1-Abs levels (Spearman r=0.35, p=0.02), the frequency of Bmems for the other DENV serotypes did not show any correlation with any of the other serotypes (Figure 6A to D). The Bmem responses did not correlate with IgG1 or IgG3 antibody responses for any of the DENV serotypes or with the age of the individuals. The frequency of Bmem cell responses to NS1 (n=43) was correlated with the frequency of neutralizing antibody titres (Neut50 titres) in all individuals (n=43) to each DENV serotype. A positive correlation was seen for DENV1 responses (Spearman r=0.50, p=0.0006) with the frequency of DENV specific Bmem responses, but no correlation was seen between responses to DENV2 (B), DENV3 (C) and DENV4 (D) (Supplementary figure 2).

**Figure 6:**
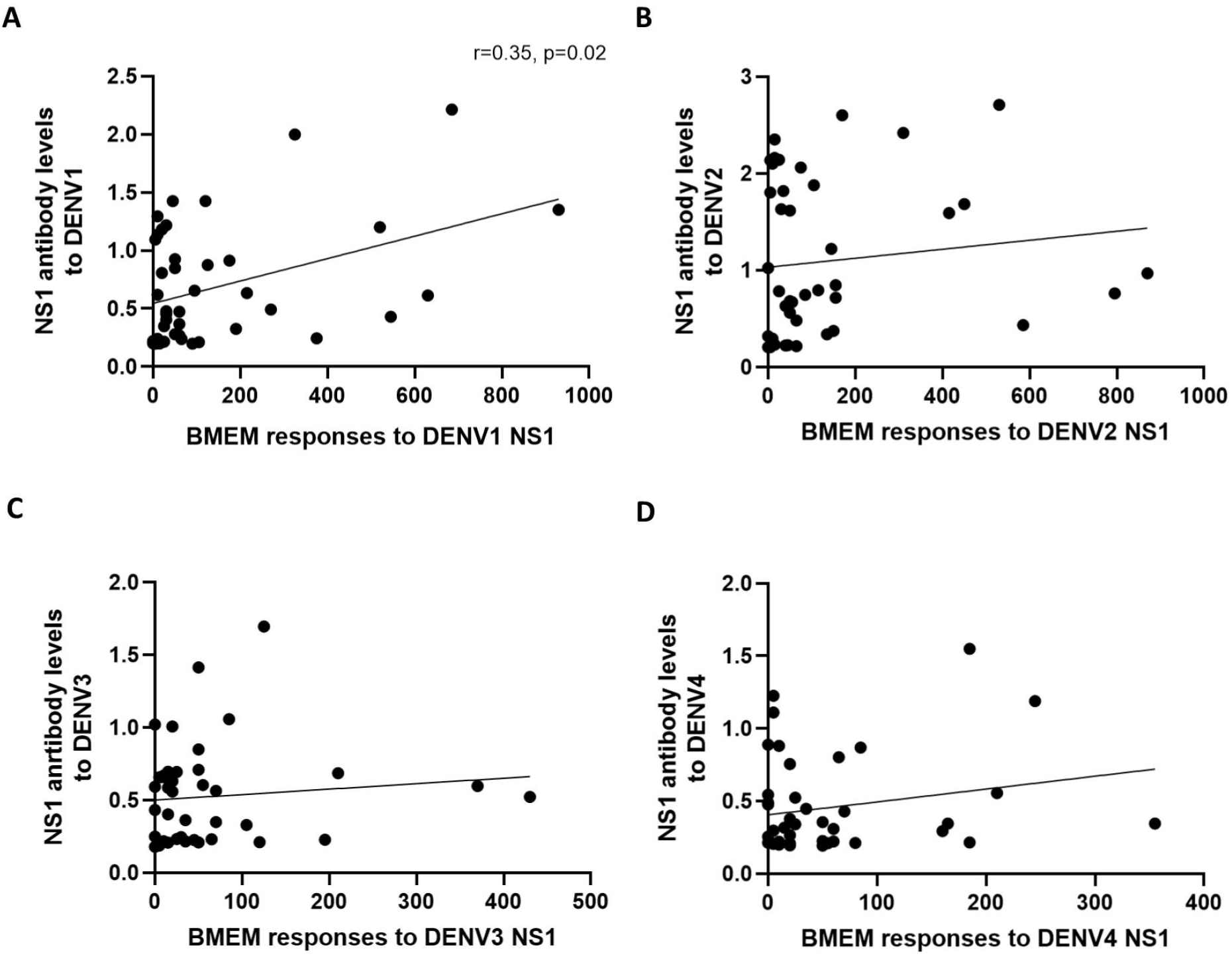
Relationship between the frequency of NS1-specific Bmem responses and NS1-specific antibody levels in individuals with varying severity of dengue infections. The frequency of NS1 specific memory B cell responses (Bmem was correlated with the frequency of NS1-specific antibody levels to each DENV serotype in all individuals (n=43) using the Spearman rank order correlation coefficient. A positive and significant correlation was seen for DENV1 responses (Spearman r=0.35, p=0.02) (A), but not between the responses for DENV2 (B), DENV3 (C) and DENV4 (D).

## Discussion

In this study we assessed the exposure to different DENV serotypes using the FRNT assay and compared the Nab levels, NS1-Ab levels, NS1-Ab levels for IgG1 and IgG3 and Bmem responses to all four DENV serotypes in individuals with varying severity of past infection. We found that there was no difference between heterotypic infections with past DF (64.29%) and DHF (68.18%), suggesting that an equal proportion of those with secondary dengue infection, experienced milder forms of dengue and DHF. In a large longitudinal study in a community cohort, it was shown that both primary and secondary dengue infections were equally likely inapparent and that the Nabs were lower in those who developed symptomatic dengue [33]. In our cohort, those with past DF had equal levels of Nabs for all four DENV serotypes, while those with past DHF had significantly higher levels to DENV1 and DENV4. Although the time of infection in those with inapparent dengue was not known, those who had DHF were infected during outbreaks of DENV1 and DENV2, where over 95% of infections were due to either serotype. Interestingly, Sri Lanka has not reported an outbreak due to DENV4 and we have not identified this serotype in hospitalised patients during our routine surveillance activities [34]. As those with past DF had broadly cross reacting Nabs, the breadth of the Nabs could influence clinical disease severity, when infected with different serotypes of the DENV.

Those with past DHF had higher NS1-Abs levels for all four DENV serotypes than in those with past DF. The higher NS1-Ab levels could be due to higher NS1 antigen levels during acute illness, which has been reported in those who progress to develop DHF [12]. Interestingly, in contrast to the observations with Nab levels, the NS1-Ab levels were highest for DENV2 in individuals with both past DF and DHF. However, in addition to the levels of antibodies, the functionality of antibodies are known to be important in protection or disease pathogenesis [32]. It was shown that those who had higher levels of total DENV specific IgG and IgG4 had inapparent dengue infection when infected with DENV3 compared to those who had lower levels [24]. Apart from neutralization of the antigen, antibodies have many functions such as complement activation, facilitating killing by natural killer cells by antibody dependent cell mediated cytotoxicity (ADCC) and phagocytosis of antibody coated cells [32]. While both IgG1 and IgG3 bind to FcγRIIa, FcγRIIIa, and FcγRIIIb, the affinity of IgG3 has shown to be higher [35]. IgG3 has shown to be critical in immune responses to many viruses and bacterial pathogens [36]. We found that those with past DHF had higher levels of NS1-specific IgG1 than individuals with past DF to three DENV serotypes and higher NS1-specific IgG3 to DENV1 and DENV4 than those with past DF. In addition, although those with past DHF had higher NS1-specific IgG1 than IgG3 to DENV1 and DENV3, there was no difference between IgG1 and IgG3 levels to any of the DENV serotypes in those with past DF. These data suggest that the NS1 specific IgG subclasses are likely to be different in those with varying severity of dengue, which would have implications for protection when re-infected with a DENV. Therefore, it would be important to further examine the functionality of NS1-specific antibody responses in those with acute illness and to carefully follow up longitudinal cohorts to determine the type of antibody repertoire that is associated with protection against severe disease.

Long lived memory B cells (Bmems) are important for recall responses to an antigen and the levels of circulating antibody levels, may not necessarily reflect the presence and frequency of Bmems for a particular antigen [37]. Although Bmems for the envelope and PrM protein of the DENV has been previously studied, NS1-specific Bmems have not been investigated before [38; 39]. We found that as seen with FRNT assays, a large proportion of our cohort had NS1-specific Bmem responses to two or more DENV serotypes. In those with past DF the highest frequencies of responses were seen to DENV2, whereas those with DHF had the highest responses to DENV1, as seen with NS1-Ab responses. However, unlike seen with NS1-Ab levels, the frequency of Bmem cells were not higher in individuals with past DHF than DF. In order to further characterise the profile of Bmem cells, it would be important to assess the IgG subclass profile and specificity of these Bmems in individuals with varying severity of dengue.

In summary, we found that those with past DF had broadly cross-reactive Nabs, while those with DHF had low levels of Nabs to some serotypes. In contrast, those with past DHF had higher NS1-Ab responses to three DENV serotypes compared to those with DF, and higher IgG1 responses. All individuals had NS1-specific Bmem responses to more than one DENV serotype. It would be important to further evaluate the functionality of NS1-specific antibody and Bmem responses to determine the type of antibody repertoire that is associated with protection against severe disease.

## Data Availability

All data is available in the manuscript and the supplementary files.

## Acknowledgements

We are grateful to the Accelerating Higher Education Expansion and Development (AHEAD) Operation of the Ministry of Higher Education funded by the World Bank, and the UK Medical Research Council.

## Figure legends

**Supplementary figure 1: Relationship between NS1-specific antibody responses and neutralizing antibody levels in individuals with varying severity of dengue infections**. NS1-specific antibody responses (n=43) positively and significantly correlated with the neutralizing antibody levels (Neut50 titres) in all individuals (n=43) to DENV1 (Spearman r=0.69, p<0.0001) (A), DENV2 (Spearman r=0.53, p=0.0003) (B), DENV3 (Spearman r=0.62, p<0.0001) (C) and DENV4 (Spearman r=0.68, p<0.0001) (D).

**Supplementary figure 2: Relationship between NS1-specific Bmem responses and neutralizing antibody levels in individuals with varying severity of dengue infections**. The frequency of memory B cell responses to NS1 (n=43) was correlated with the frequency of neutralizing antibody titres (Neut50 titres) in all individuals (n=43) to each DENV serotype. A positive correlation was seen for DENV1 responses (Spearman r=0.50, p=0.0006) (A), with no correlation between responses to DENV2 (B), DENV3 (C) and DENV4 (D).

