## Supplementary figures and images for "Antibody and memory B cell responses to the dengue virus NS1 antigen in individuals with varying severity of past infection"

### Supplementary fig 1

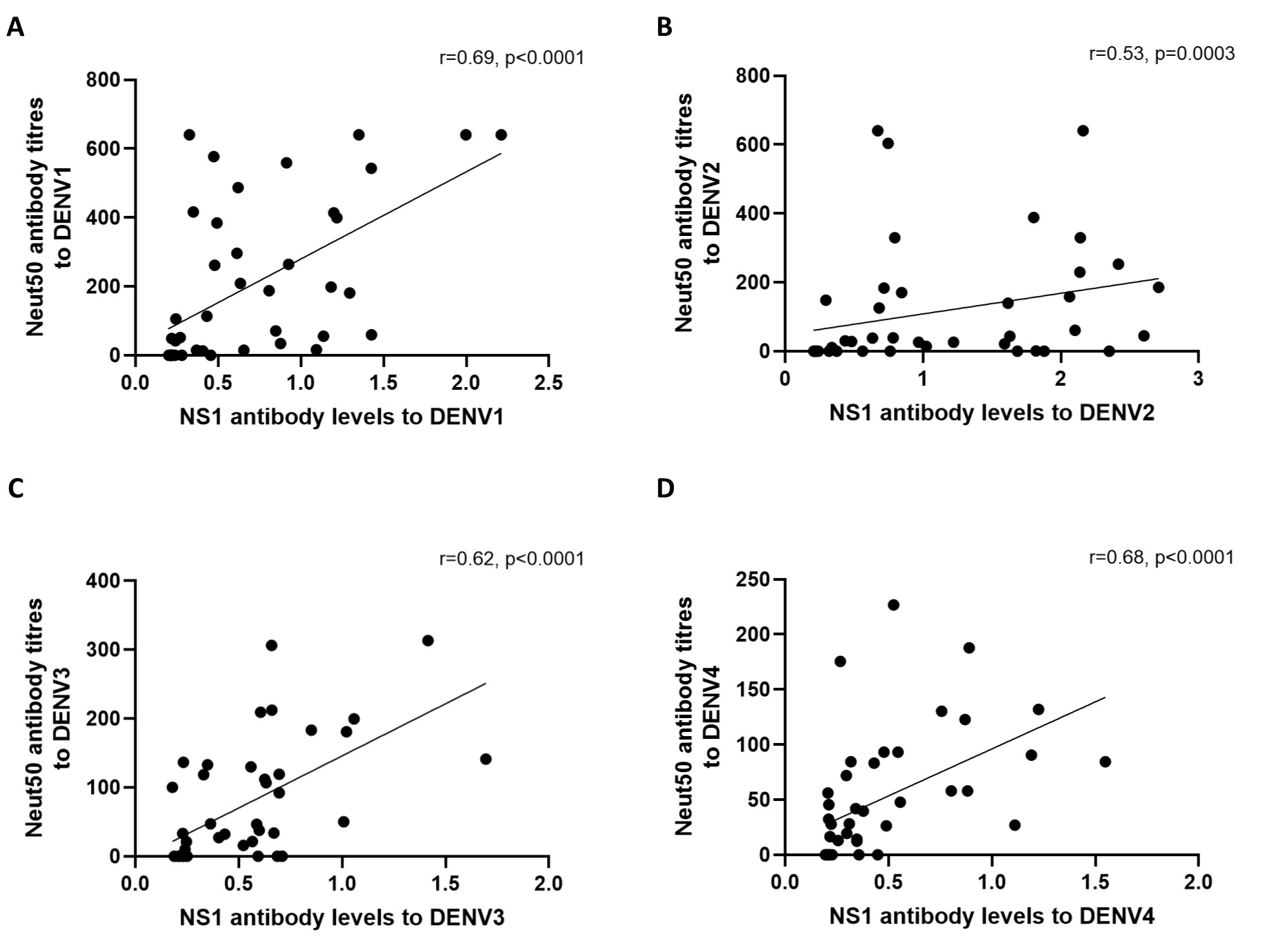

### Supplementary fig 2

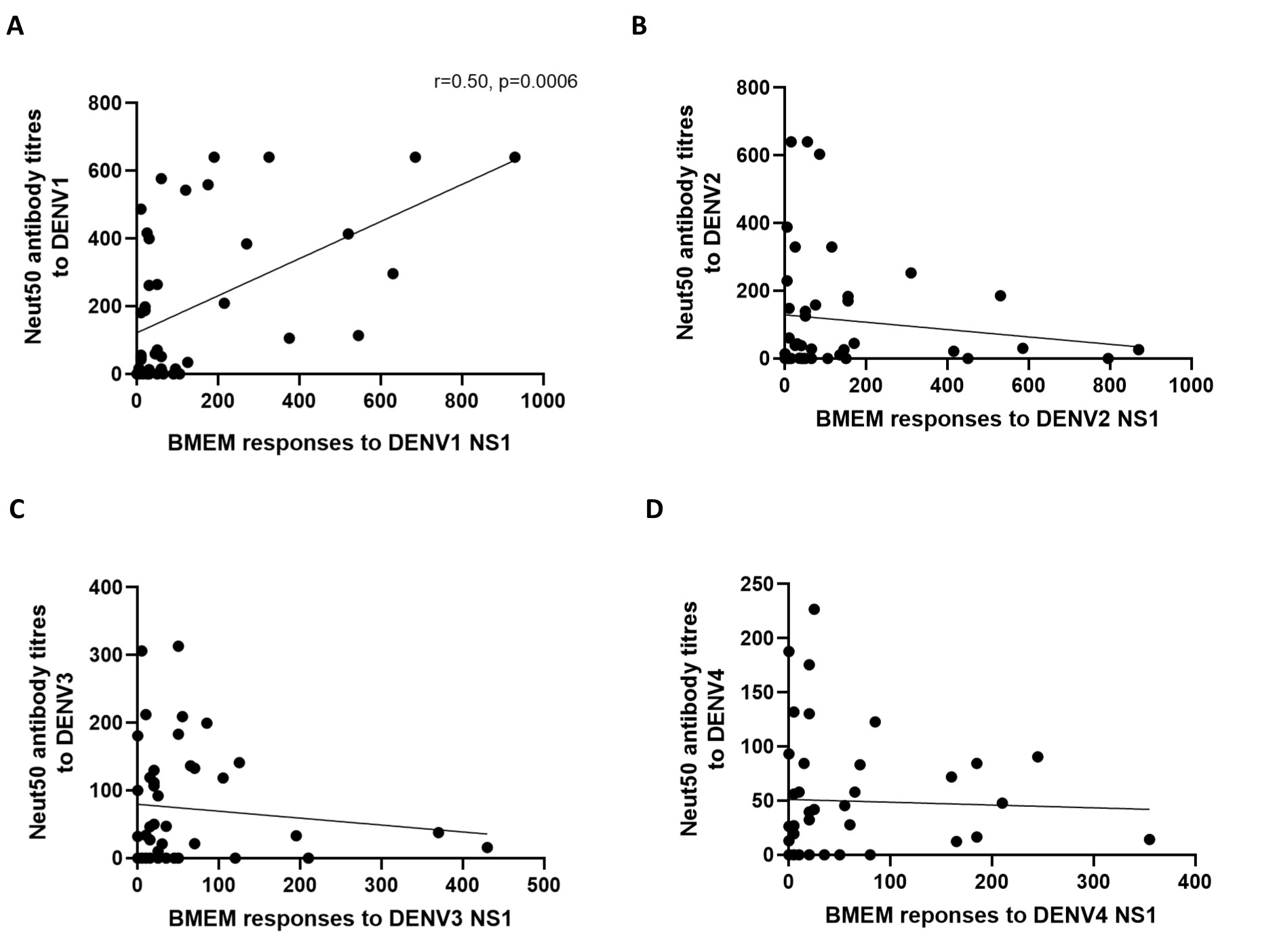
